# Risk of SARS-CoV-2 reinfections in children: prospective national surveillance, January 2020 to July 2021, England

**DOI:** 10.1101/2021.12.10.21267372

**Authors:** Anna A Mensah, Helen Campbell, Julia Stowe, Giulia Seghezzo, Ruth Simmons, Joanne Lacy, Antoaneta Bukasa, Shennae O’Boyle, Mary E Ramsay, Kevin Brown, Shamez Ladhani

## Abstract

**Background:** Reinfection after primary SARS-CoV-2 infection is uncommon in adults, but little is known about the risks, characteristics, severity or outcomes of reinfection in children.

**Methods:** We used national SARS-CoV-2 testing data in England to estimate the risk of reinfection ≥90 days after primary infection from 01 January 2020 to 31 July 2021, which encompassed both the Alpha and Delta waves in England. Disease severity was assessed by linking reinfection cases to national hospitalisation, intensive care admission and death registrations datasets.

**Findings:** Reinfection rates closely followed community infection rates, with a small peak during the Alpha wave and a larger peak during the Delta wave. In children aged ≤16 years, there were 688,418 primary infections and 2,343 reinfections. The overall reinfection rate was 66·88/100,000 population, being higher in adults (72.53/100,000) than in children (21·53/100,000). Reinfection rates after primary infection were 0·68% overall, 0·73% in adults and 0·34% in children. Of the 109 reinfections in children admitted to hospital, 78 (72%) had underlying comorbidities. Hospitalisation rates were similar for the first (64/2343, 2·73%) and second episode (57/2343, 2·43%). Intensive care admission was rare after primary infection (n=7) or reinfection (n=4), mainly in children with comorbidities. 44 deaths occurred after primary infection within 28 days of diagnosis (44/688,418, 0·01%), none after possible reinfections.

**Interpretation:** The risk of SARS-CoV-2 reinfection is strongly related to exposure due to community infection rates, especially during the Delta variant wave. Children had a lower risk of reinfection than adults, but reinfections were not associated with more severe disease or fatal outcomes.

**Funding:** PHE/UKHSA

**Research in Context:** *Evidence Before this study:* We searched PubMed with the terms “COVID-19” or “SARS-CoV-2” with “reinfection” to identify publications relating to SARS-CoV-2 reinfections from 01 January until 15 November 2021. There were few publications relating to SARS-CoV-2 reinfections, and these primarily related to adults. Published studies reported very low rates of reinfection during the first few months after primary infection in adults. COVID-19 vaccines provide effective immune protection against SARS-CoV-2 infection, but recent studies have reported increasing risk of breakthrough infection with time since primary vaccination due to waning immunity. Several SARS-CoV-2 variants, including the beta, gamma and delta variants have been shown to partially evade immunity after natural infection and vaccination, potentially increasing the risk of reinfections and breakthrough infections, respectively. Data on reinfections in children are lacking and restricted mainly to case reports in immunocompromised children.

*Added Value of This Study:* We used national SARS-CoV-2 testing data during the first 19 months of the pandemic to estimate the risk of reinfection in children compared to adults during a period that encompassed both the Alpha and the Delta variant waves in England. We found that the risk of reinfection correlated with the risk of SARS-CoV-2 exposure and therefore, closely reflected community infection rates, with most reinfections occurring during the Delta variant wave. Whilst acknowledging the limitation of using national testing data, we found that children had a lower risk of reinfection compared to adults and that the risk of reinfection in children increased with age. Reinfections were not associated with severe disease in terms of hospitalization or intensive care admission and there were no fatalities within 28 days of the reinfection episode in children.

*Implications of all the Available Evidence:* SARS-CoV-2 reinfections are rare in children, especially younger children, and occurred mainly during the Delta wave in England. Reinfections were not associated with more severe disease or fatal outcomes in children. COVID-19 vaccination will provide further protection against primary infections and reinfections in children.

## Introduction

Children have been relatively spared by SARS-CoV-2 in the COVID-19 pandemic. When exposed to SARS-CoV-2, children are more likely than adults to develop a mild, transient illness and less likely to develop severe disease, require hospitalisation or intensive care admission, or die of COVID-19.^1^ Following infection, children develop robust humoral and cellular responses to SARS-CoV-2, targeted primarily against the spike protein of the virus, irrespective of whether they develop symptomatic illness or remain asymptomatic after infection.^2^ Additionally, immunity is sustained for at least 12 months, potentially better than in adults, with evidence of some cross-protection against new SARS-CoV-2 variants.^2^ Reinfection following primary SARS-CoV-2 infection is uncommon but has been described mainly in adults, even in the presence of SARS-CoV-2 antibodies.^3-5^ Additionally, breakthrough infections have been reported in vaccinated adults, with increasing risk with time since vaccination.^5^ An increased risk of reinfection has also been reported following the emergence of the Delta (B.1.617.2) variant, which is more transmissible than previous SARS-CoV-2 variants and more able to at least partially evade both natural and vaccine-induced immunity.^6^ However, little is known about the rate, risk factors, characteristics, severity or outcomes of SARS-CoV-2 reinfections in children with the exception of a few cases reports in immunocompromised children.^7,8^

In England, the first cases of SARS-CoV-2 emerged in late January 2020 and increased rapidly, leading to national lockdown including school closures in March 2020.^9^ Cases peaked in mid-April and then declined gradually, leading to gradual easing of lockdown measures, including partial reopening of some school class years during June/July 2020, and full reopening of all school years albeit with strict infection controls in September 2020.^10^ Early in the pandemic, testing for SARS-CoV-2 was limited to symptomatic cases in healthcare settings, but community testing became available from June 2020, initially for individuals with typical symptoms of COVID-19 (fever, new onset cough or loss of taste or smell) but soon became available for anyone wishing to be tested for whatever reason, with a 7-day rolling average of in excess of 100,000 tests per day by 28 June 2020.^11^

In order to assess the risk of SARS-CoV-2 reinfection in children compared to adults, we analysed national testing data for England during the first 19 months since the start of the pandemic, which included the Alpha variant wave during the winter of 2020/2021 and the Delta variant wave during summer 2021.

## Methods

The UK Health Security Agency (UKHSA) and its predecessor, Public Health England, collects and compiles all SARS-CoV-2 tests performed nationally through community and healthcare settings using the electronic Second Generation Surveillance System (SGSS), which includes information on patient’s sex, age and the region the test was performed. We obtained results for all positive SARS-CoV-2 diagnostic tests performed in England from 27 January 2020 until 31 July 2021. Primary or first infections were defined as the first ever positive SARS-CoV-2 RT-PCR or rapid antigen or lateral flow device (LFD) test result for an individual. A possible reinfection was defined as a subsequent positive SARS-CoV-2 RT-PCR or LFD test result at least 90 days from the previous positive test. Individuals testing positive for SARS-CoV-2 by LFD testing are advised to take a confirmatory RT-PCR test. Consequently, those who subsequently tested PCR negative within 3 days were removed from the dataset listing all possible reinfections. If an individual tested positive for SARS-CoV-2 on multiple occasions with a minimum interval of 90 days between each episode, then only the first and second episodes were included in the analysis.

We used the Secondary Uses Service (SUS), which is a national electronic database of hospital admissions that provides timely updates of ICD-10 codes for completed hospital stays for all NHS hospitals in England, to identify children (0-16 year-olds) attending hospital and/or an intensive care unit (ICU) within 21 days of testing positive for SARS-CoV-2. We also used ICD-10 codes recorded for primary and secondary diagnoses in SUS to identify children with comorbidities. Finally, we linked cases to electronic deaths registrations provided by the Office for National Statistics (ONS) to identify children who died (i) within 28 days of testing positive for SARS-Cov-2 or (ii) within 60 days and/or with COVID-19 recorded on their death certificate.

### Data analysis

We calculated rates of primary infection and reinfection per 100,000 population using the ONS 2020 mid-year estimates for population denominators.^12^ We compared numbers and rates of primary infection and reinfections by age in years in children and by age-group (<5 years, 5-11 years or primary school-aged and 12-16 years or secondary school-aged) and compared rates with adults aged 17 years and over, who were eligible for COVID-19 vaccination from December 2020, in a structured rollout that started with 80+ year-olds along with health and social care workers and then extended down the age groups and those with specific underlying comorbidities.^13^ COVID-19 vaccine uptake by age and time was obtained from the online NHS England COVID-19 Vaccination archive.^14^

## Results

### Primary infections and reinfections

During the surveillance period, primary infection cases were low during the summer of 2020, then increased from the end of August just before the start of the Autumn school term and peaked at the end of December 2020 following the emergence and rapid spread of the Alpha variant across England since mid-November 2020 (**Figure 1**). A national lockdown, including school closures, in January 2021 was associated with a rapid decline in cases, with a small peak in March 2021 when children returned to school whilst adults remained in national lockdown, followed by a larger peak from mid-May 2021 with the easing of national lockdown alongside the emergence and rapid spread of the Delta variant nationally.

**Figure 1:**
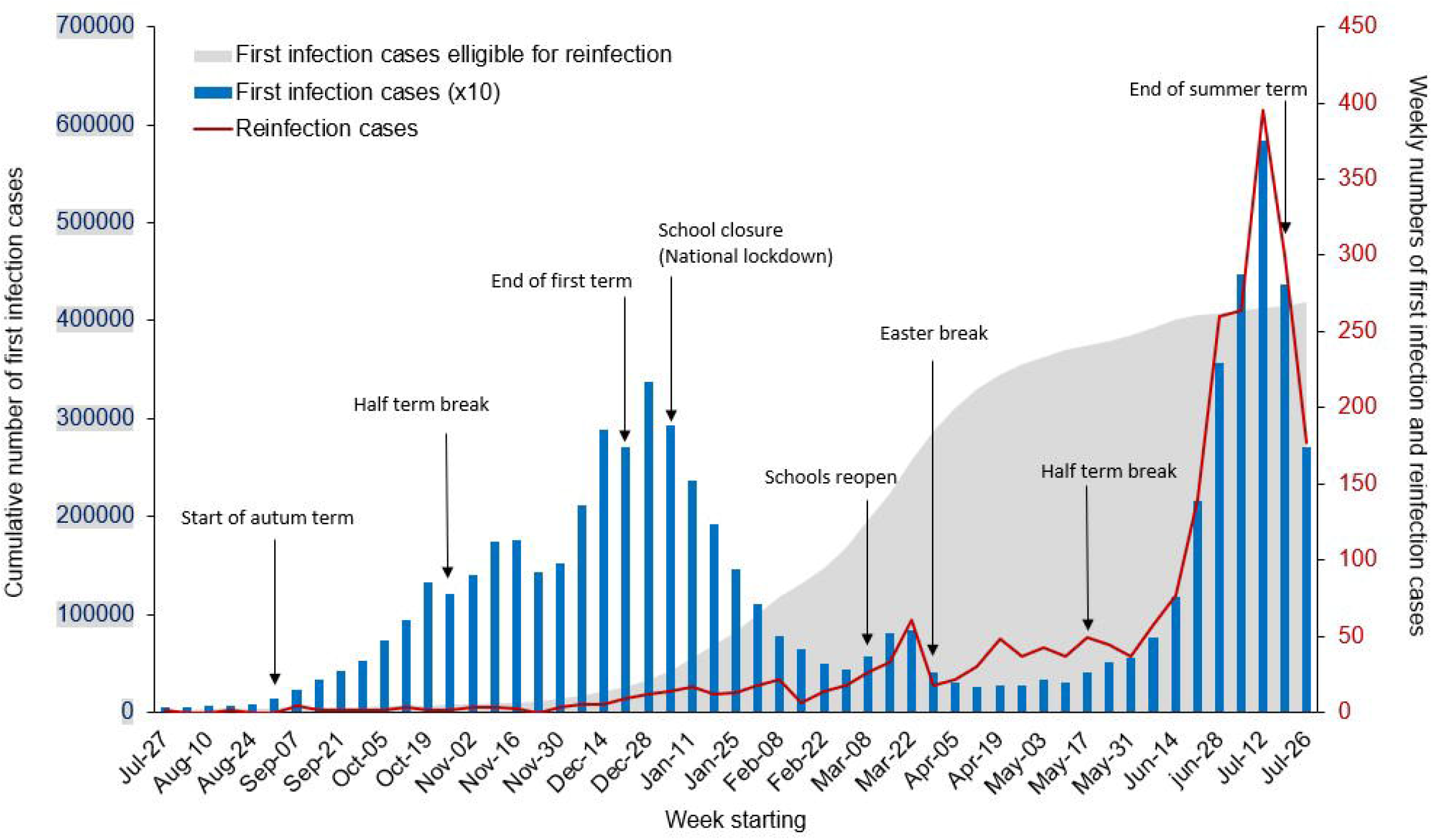
Weekly cumulative laboratory-confirmed SARS-CoV-2 primary infection cases eligible for reinfection (allowing for 90 days interval), weekly primary infection and weekly numbers reinfection cases in children aged 0-16 years between 27 July 2020 and the week beginning 26 July 2021 in England. Note that the numbers of weekly first infection cases have been multiplied by ten to allow visual comparison with the reinfection cases.

Reinfection rates were very low in children during the Alpha wave, at a time when the number of primary infection cases eligible for reinfections was also low. There was a small increase in reinfection cases during March 2021 as the cumulative count of primary cases eligible for reinfection increased rapidly by almost 6-fold compared to the number of cumulative cases during the winter peak in December (**Figure 1**). Reinfection cases then stabilised for two months before increasing rapidly from mid-May 2021 and following the same trajectory as community infection rates during the Delta wave, reaching a peak at the same time as primary infection rates in mid-July 2020. Weekly primary infection and possible reinfection cases subsequently declined after schools closed for the summer holidays.

When assessed by age-group, children were the only age-group that had not exceeded a weekly reinfection rate of 1 per 100,000 individuals until the week starting 14^th^ June 2021 (**Figure 2**). During the Alpha wave, reinfections occurred mainly in older adults (80+ year olds). As the adult population became increasingly vaccinated since December 2020 (**Supplement Figure 1**), the age distribution of reinfection cases changed, with the emergence of the Delta variant, the reinfection risk was highest in the younger adults (17-39 year-olds), who remained largely unvaccinated or partially vaccinated with a single vaccine dose. During the Delta wave, reinfection rates in 12-16 year-olds peaked in the week beginning 12 July 2021 and mirrored reinfection rates in 50-59 year-olds who had been 93% and 73% vaccinated with their first and second COVID-19 doses, respectively, by 01 June 2021 (**Supplement Figure 1**).

**Figure 2:**
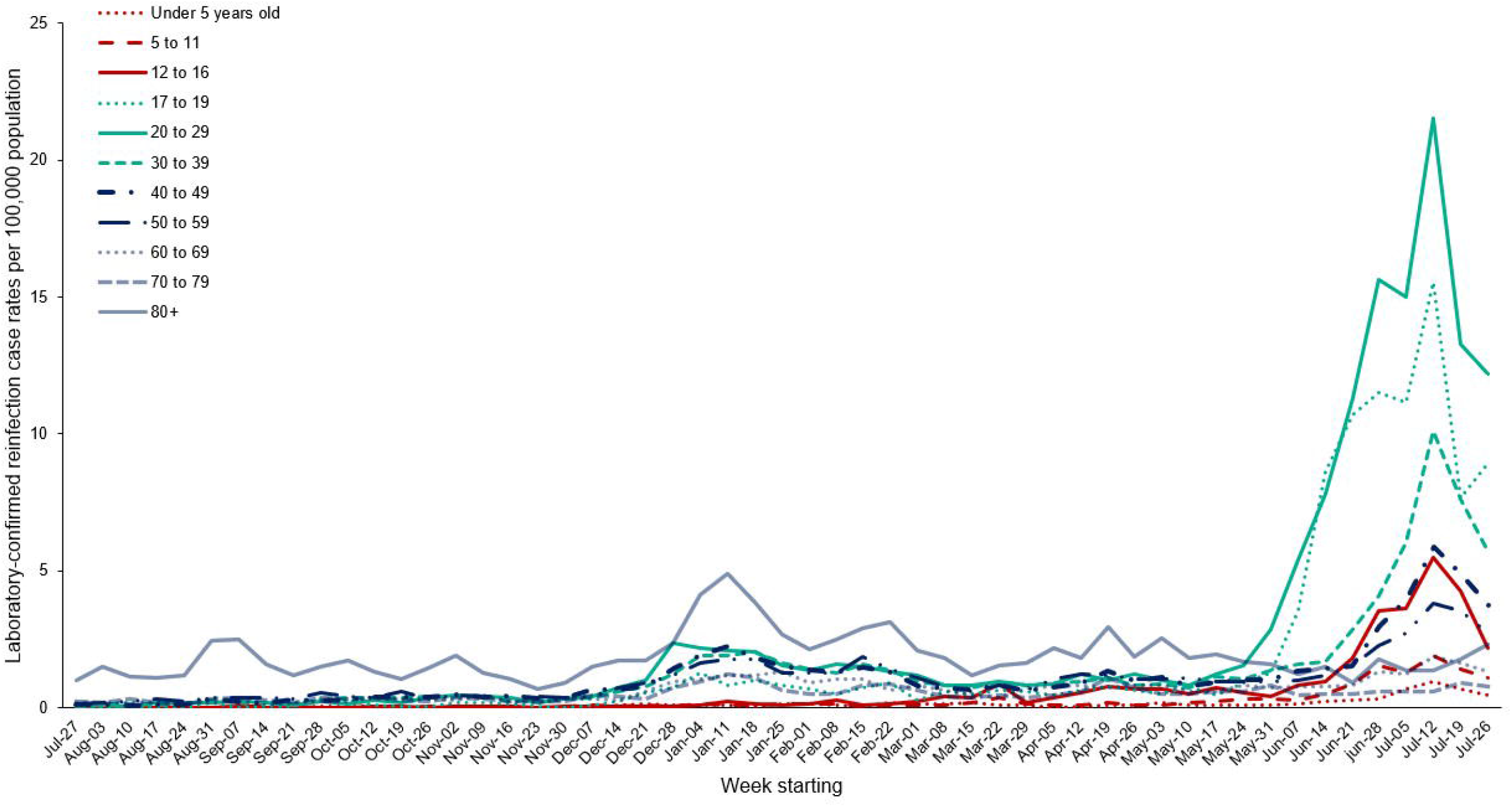
Weekly rates of laboratory-confirmed SARS-CoV-2 reinfection cases per 100,000 population be age group in children and adults between 27 July 2020 and the week beginning 26 July 2021, England

In children, at the peak of reinfections in the week beginning 12 July 2021, there was an age-related gradient, with the lowest reinfection rate of 0·9 per 100,000 in <5 year-olds compared to 1·9 per 100, 5-11 year-olds and 5·5 per 100,000 in 12-16-year-olds. These rates were 23, 11 and 4 times lower than in adults aged 20-29 years, who were unvaccinated and with the highest reinfection rate at the time (**Figure 2**).

During the 19-month surveillance period (January 2020 to July 2021), primary childhood infection rates were higher in infants (<1 year-olds) and then dropped in 1 and 2 year-olds before increasing with age up to 16 years (**Figure 3**). Reinfections, on the other hand, were lowest in infants (23 cases) and remained low among 1-6 year-olds with an average of 43 cases per year group, before increasing with age, reaching 364 cases among 16 year-olds.

**Figure 3:**
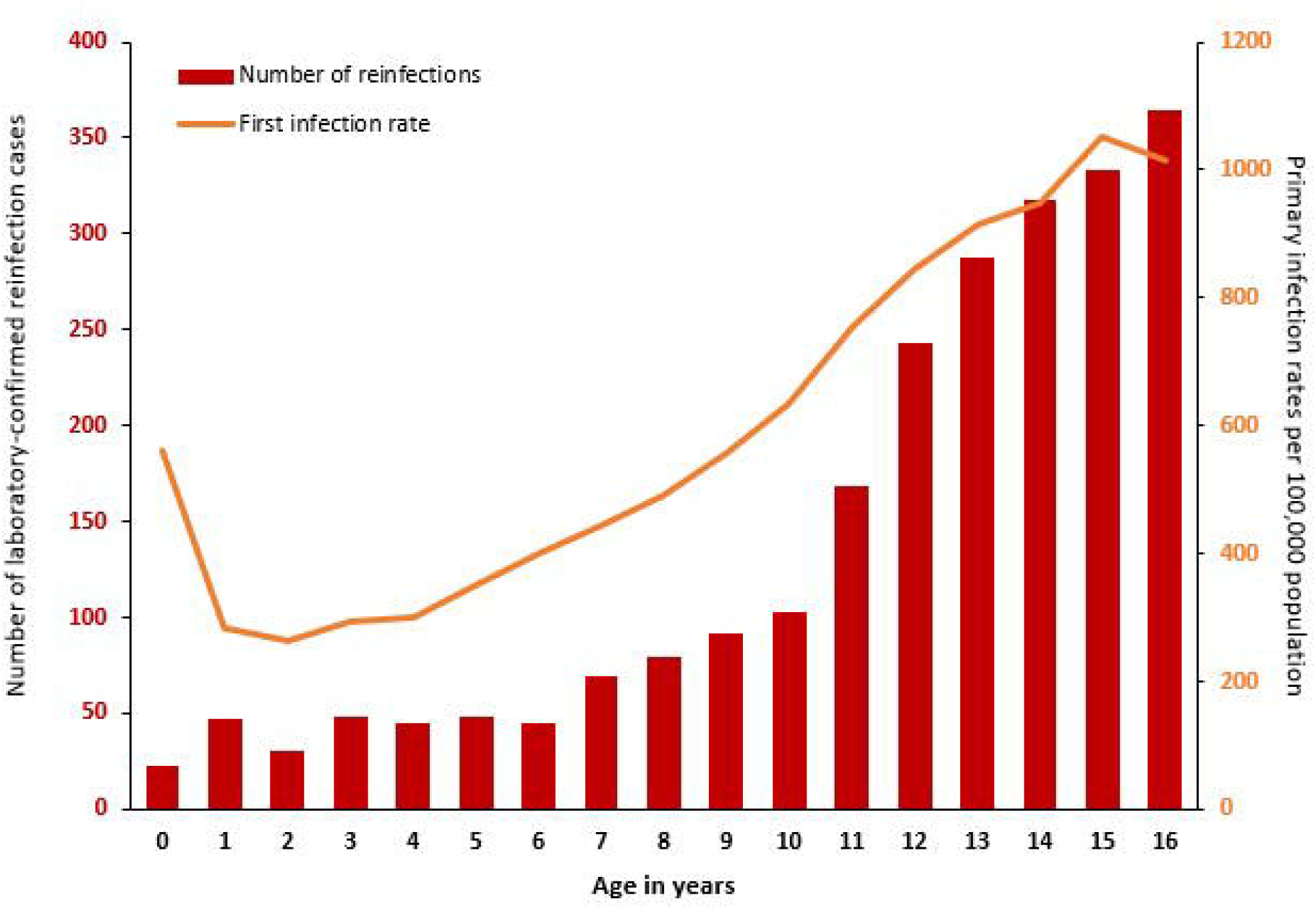
Laboratory-confirmed SARS-CoV-2 reinfection cases by year of age in children (primary y-axis) and incidence of primary SARS-CoV-2 infections in children aged 0-16 years between 01 January 2020 until 31 July 2021, England

### Reinfection rates

In England, primary infections in children started increasing in February 2020 with the first reinfection in a child identified in late July 2020. By end July 2021, there were 688,418 primary infections and 2,343 possible reinfections in children. There was no gender (341,981/688,418 [50·14%] vs. 1,160/2,334 [49·70%] in males where this information was available) or ethnicity differences (458,622/619,388 [74·04%] vs 1,477/1,973 [74·86%] in people of white ethnicity where information was available) between primary and secondary infections. The overall reinfection rate for this period was 66·88/100,000 population, being higher in adults (72·53/100,000) than in children (21.53/100/000). Reinfection rates after primary infection over the same period were 682/100,000 primary infections overall, 730/100,00 in adults and 343/100,000 in children. Reinfection rates by age-group are summarised in **Figure 4**.

**Figure 4:**
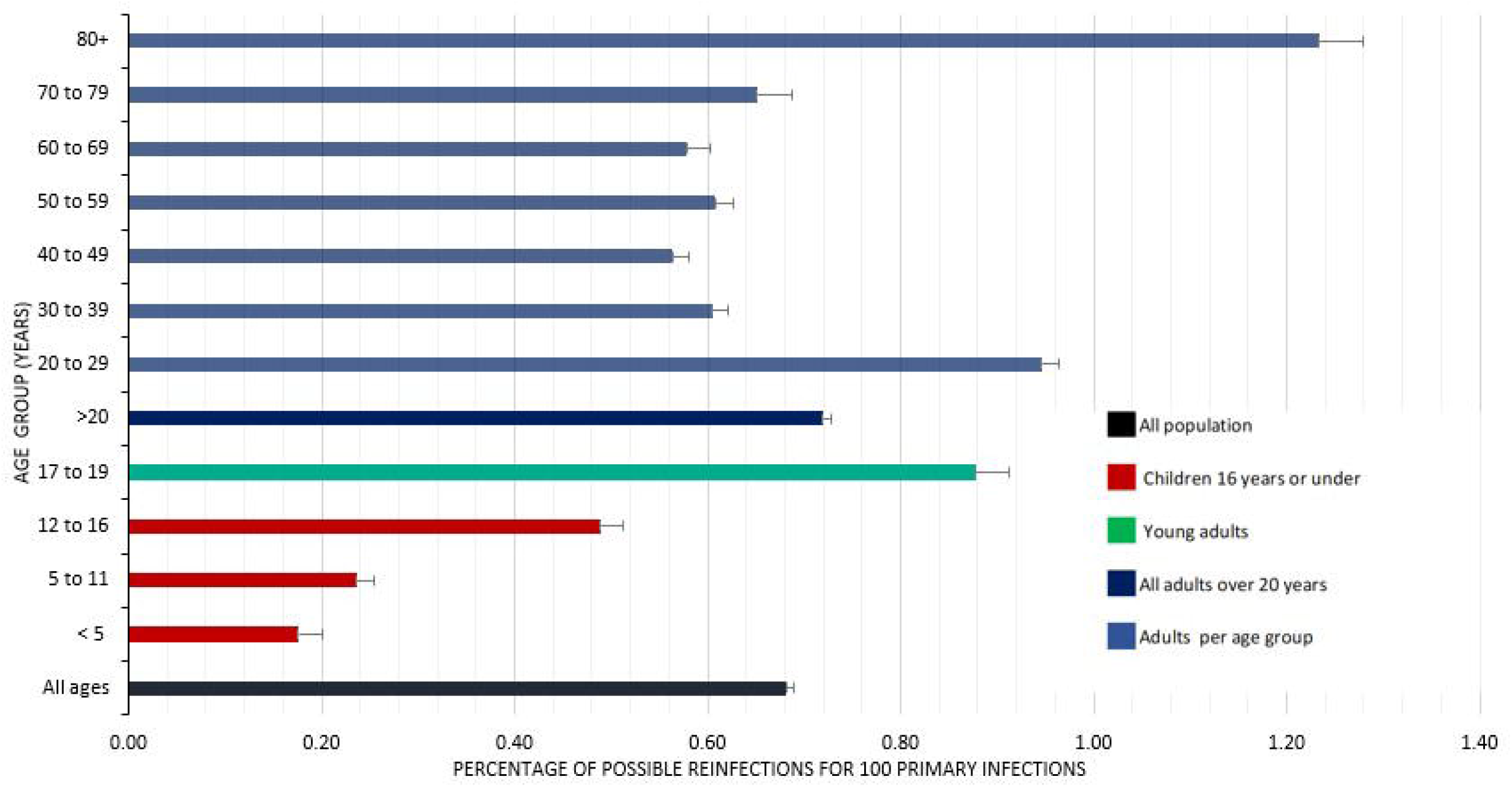
Laboratory-confirmed reinfection rates with 95% confidence interval by age-group for the period January 2020 to 31 July 2021, England

### Disease severity and outcomes

Of the 2,343 children with reinfection, 109 (4·65%) were admitted to hospital at either episode and 78 (72%) of those hospitalised were identified as having an underlying comorbidity. Hospitalisation rates in children with reinfection were similar for the first (64/2343, 2·73%) and second episode (57/2343, 2·43%) and included 12 children who were hospitalised for both episodes that all had additional health issues.

While reviewing the ICD-10 codes provided with hospital admission, 77% (49/64) and 72% (41/57) of children admitted to hospital had other codes unrelated to COVID-19 diagnosis recorded at first and second episode respectively. In addition, 5 children at each episode were admitted to hospital specifically for injuries.

Median (IQR) age of children was slightly lower at 11 years old (IQR=7-14) for primary infection cases compared to 13 years old (IQR= 10-15) for possible reinfection cases.

The proportion of children reporting asymptomatic infection at the time of testing was higher during the second episode compared to the primary episode (1,038/2,113 [49%] vs 235,914/638,389 [37%]; P<0.0001)

Admission to an intensive care unit was rare after primary infection (n=7) or reinfection (n=4). Notably, all 4 children admitted to an intensive care unit following reinfection had also required intensive care during their primary infection. All 4 children had multiple and severe multi-system co-morbidities and, despite detailed case note review, it was not possible to ascertain the contribution of SARS-CoV-2 infection to the illness that eventually led to the intensive care admission.

There were 44 deaths within 28 days of testing positive for SARS-CoV-2 (44/688,418 primary infections, 0·01%), with a further 9 deaths occurring within 60 days of testing positive or with COVID-19 specified as a cause of death on the death certificate (53/688,418 primary infections, 0·01%). All deaths occurred following primary infection and there were no deaths within 28 days of positive testing for SARS-CoV-2 after reinfection.

## DISCUSSION

SARS-CoV-2 reinfections in children were uncommon and correlated closely with community infection rates in England, with the majority of reinfections occurring during the Delta wave in summer 2021. Children had a lower risk of reinfection compared to adults overall, especially when compared to unvaccinated younger adults. Among children, reinfections were low up to 6 years of age and then increased steadily up to 16 years of age, broadly consistent with their risk of primary infection. Children experiencing severe infections, as evidenced by hospitalisations or intensive care admissions, were rare following both primary infection or reinfection, with a high prevalence of comorbidities among hospitalised children with reinfection and no deaths reported up to 28 days after reinfection episode.

There are several important factors that need to be considered when assessing the risk of reinfection in this cohort. Firstly, reinfection cases were based on sequential positive tests with an interval of ≥90 days alone and did not require a negative PCR result between infection episodes or a genetically distinct SARS-CoV-2 strain at each episode, as specified in other definitions.^3^ As of 31 October 2021, there were only 441 confirmed reinfections with genetically distinct strains in England.^15^ We used a pragmatic definition applied to surveillance data that allowed sufficient time for clinical recovery and for the initial PCR test to become negative before a second infection was diagnosed to rule out persistent infections, whilst acknowledging that reinfection may rarely occur within 90 days of primary infection.

Secondly, there was limited testing for SARS-CoV-2 early in the pandemic, with testing restricted primarily to symptomatic individuals attending healthcare settings. Since children are significantly less likely than adults to develop symptomatic infection let alone severe disease or be hospitalised for COVID-19, it is likely that earlier infections would not have been laboratory-confirmed. Our serosurveillance in educational settings estimated that 11·2% of primary school students (5-11 year-olds) had been infected by SARS-CoV-2 by June 2020^16^ and 12·8% of secondary school students by September 2020 (11-16 year-olds).^17^ Thirdly, community testing became more widely available from June 2020, including for children, but was primarily targeted for symptomatic individuals with typical COVID-19 symptoms (fever, new-onset cough and loss of taste or smell). Since children are more likely than adults to develop a mild, transient illness when exposed to SARS-CoV-2 (and, therefore, not seek medical attention for testing or treatment) and also to develop non-respiratory symptoms,^18^ they would potentially be less likely than adults to get tested even after community testing was available. Although, with increasing community testing capacity, everyone had access to testing for any reason and not restricted to testing after typical COVID-19 symptoms, as evidenced by the high proportion of asymptomatic individuals at the time of testing. Fourthly, national testing data would not pick up asymptomatic primary infections or reinfections unless individuals got tested because they were concerned over exposure to a case, and children are more likely to be asymptomatic – and, therefore, less likely to be tested - than adults.^19^ Fifthly, the introduction of twice-weekly rapid home testing with LFDs for secondary schools since March 2021 will likely have enhanced case ascertainment, including asymptomatic infections, leading to reduced virus introduction and in-school transmission in 11-16 year-olds. Finally, the risk of reinfection is also likely to vary with time since primary infection and with different SARS-CoV-2 variants. Comparison of relative risks between variants, however, is not possible because they emerged at different times, such that both the number of primary infections who became susceptible to reinfection and the interval between primary infection and reinfection were far greater during the Delta wave in July 2021 compared to the Alpha wave in December 2020.

Furthermore, comparison with adults is hampered by differential exposure risks over time, with prolonged periods of national lockdown, which at times included school closures. Schools remained open for in-person teaching during national lockdown in June/July 2020, November 2020 and March-May 2021, for example, but closed during January/February 2021 when England was experiencing the second pandemic wave due to the Alpha variant.^20^ Among school-aged children, too, testing rates were heavily influenced by school closure either due to lockdowns or school term breaks.^21^ The national rollout of COVID-19 vaccines from December 2020, with gradual extension of eligibility down the age groups, also affected comparison of risk and rates over time.

Whilst keeping these considerations in mind, we have for the first time estimated the risk of reinfections in children and compared them to adults over a 19-month period. Reinfections occur frequently after infection with seasonal coronaviruses which cause the common cold (229E, OC43, NL63, and HKU1) because of short-lasting, poorly cross-protective immunity between infections.^22^ Reinfections have also been reported with SARS-CoV-2, although the risk was previously estimated to be very low during the first few months after primary infection in adults and even lower in children, although there are very limited data on reinfections in the paediatric population.^3,23,24^ Our study confirms a lower risk of reinfection in children compared to adults, especially unvaccinated young adults and, within the childhood age-groups, reinfection rates broadly correlated with primary infection rates by age in years except in infants (<1 year-olds) who had a higher primary infection rate compared to reinfection rate. This is likely to be a testing artefact because of new-born screening for SARS-coV-2 after birth and as part of infection screens in young infants presenting to hospital with fever and/or an infectious illness.^25^

Whilst the vast majority of individuals develop robust protection after SARS-CoV-2 exposure, there is wide heterogeneity in the immune response after primary infection, in terms of neutralising antibodies and T and B cell repertoire, which may render a small proportion of individuals susceptible to reinfection, especially those who were asymptomatic or had mild illness with their primary infection.^3^ Reinfections may also occur because of waning immunity with time since primary infection; whilst most children and adults retain high antibody levels with virus neutralising activity for at least 12 months after primary infection, they do wane to undetectable levels in a small proportion.^2^ Waning of immunity has also been reported among vaccinated adults, resulting in increasing risk of breakthrough infections with time since vaccination,^26^ although this risk appears to be lower in mRNA-vaccinated adults with prior SARS-CoV-2 infection compared to vaccinated adults without prior infection.^27^

Importantly, too, the risk of reinfection appears to be associated with emergence of new variants - such as the Beta, Gamma and Delta variants - which can at least partially evade immunity from prior infection as well as vaccination.^5^ In our cohort, reinfection rates were much higher during the Delta wave compared to the Alpha wave, which may be because the Alpha variant is antigenically similar to previously circulating SARS-CoV-2 strains whilst the Delta variant, like the Beta variant, are antigenically distant.^28^ This is consistent with the lower neutralising activity of antibodies after natural infection,^28^ or vaccination,^29,30^ as well as lower vaccine effectiveness after mRNA vaccination,^31^ against the Delta variant compared to the Alpha and previous variants.

Reassuringly, though, prior infection or COVID-19 vaccination and, more so, a combination of the two have shown to be highly protective against severe disease and death due to COVID-19, irrespective of the responsible variant, with studies reporting higher rates of asymptomatic infection and mild illness following reinfection compared to the primary infection in the same individuals.^3,4,31^ This was also observed in our cohort of children who were more likely to be asymptomatic with their reinfection compared to primary infections. Among children with reinfection, the risk of hospitalisation is similar to overall hospitalisation rates for confirmed childhood COVID-19,^32^ whilst ICU admissions were rare and mainly in those with severe underlying co-morbidities. In adults, underlying comorbidity is an important risk factor for reinfections.^4^ Lastly, further work is needed to assess the risk of long covid following reinfection compared to primary infection as the persistence of a wide array of symptoms lasting months after SARS-CoV-2 infection is a non-negligeable part of the disease burden elicited by COVID-19.

### Limitations

In addition to the limitations already discussed, we did not collect detailed clinical data on all the reinfection cases to assess disease severity compared to the primary infection. Among hospitalised cases, too, we were unable to distinguish whether the children were hospitalised for COVID-19 or the virus was identified incidentally during routine screening of children attending hospital for other illnesses. The emergence of new variants also made it difficult to assess waning of natural immunity over time. As children continue to be vaccinated against COVID-19, it will become more difficult to assess long-term protection after natural infection.

## Conclusions

In children, the risk of SARS-CoV-2 reinfection is strongly related to the risk of exposure and, therefore, community infection rates. Reinfection rates were higher during the Delta wave than the Alpha wave, most likely the result of a combination of waning immunity over time since primary infection and partial immune evasion by the Delta variant compared to the Alpha variant. In children, the risk of reinfection increased with age. The vast majority of children had mild infection with very low hospitalisation rates after primary infection or reinfection.

## Supporting information

Ethics approval

## Data Availability

All data produced in the present study are available upon reasonable request. Requests to access non-publicly available data are handled by the PHE now UKHSA Office for Data Release (ODR).

## Contributors

AAM, HC, SL: methodology; formal analysis; investigation; data curation; writing the original draft; writing – review and editing; visualisation

JS, GS, RS, SOB, AB, JL: data curation; writing-review and editing

SL, HC, KB: conceptualisation; methodology; supervision; writing – review and editing

## Legends

**Supplemental figure 1:**
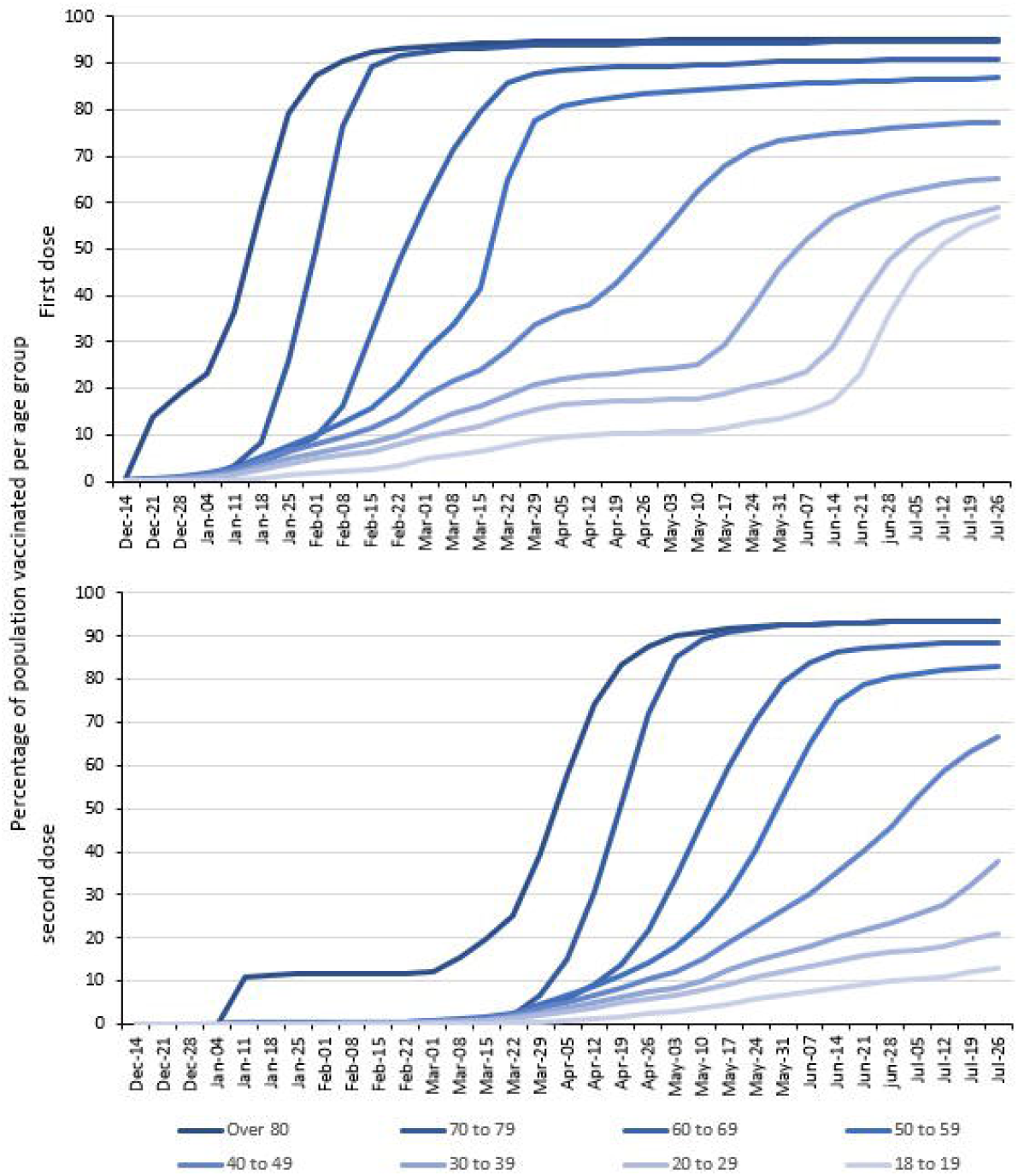
Cumulative weekly COVID-19 vaccination uptake in by age group and over time, England. Children were not eligible for COVID-19 vaccination during the surveillance period.

