## Supplementary material for "Risk of SARS-CoV-2 reinfections in children: prospective national surveillance, January 2020 to July 2021, England": Ethics approval

Public Health England

PHE Research Support and Governance Office
155 Waterloo Road, London SE1 8UG

www.phe.gov.uk

18th February 2021

Dr Helen Campbell
Senior Clinical Scientist
PHE
Immunisation and Countermeasures Dept
National Infection Service
61 Colindale Avenue
London
NW9 5EQ

Dear Dr Campbell

**Re:** National surveillance of possible symptomatic COVID-19 re-infection

**R&D Ref:** NR0259

Thank you for submitting your study to the Research Support and Governance Office (RSGO) for review by the PHE Research Ethics and Governance Group (REGG).

PHE approval for your study has been granted and you can now start your work in this organisation. This approval is granted based on the information provided in the REGG application form and accompanying study documentation, and on the understanding that the study is conducted in accordance with the conditions stated in the applicable PHE policies and procedures.

Approval is only granted for activities for which a favourable opinion has been given by the PHE REGG. All amendments must be submitted to the RSGO. Any change to the status of the project (including changes to the research team) must also be notified to the RSGO.

PHE is currently undertaking the implementation of an end to end research management system and institutional repository. Aligned to this, from 1 September 2020 the PHE Open Access policy requires peer-reviewed research outputs to be made available open access. For further information contact Paul Rudd.

If you need any further support or information, please do not hesitate to contact the PHE RSGO quoting the reference number for your study.

Wishing you every success with your study

Yours sincerely,

E Coates

Dr Elizabeth Coates
Head of Research Governance
Research Support and Governance Office
